# Maternal thyroid hormone imbalance and risk of autism spectrum disorder

**DOI:** 10.1101/2025.01.07.25320099

**Authors:** Leena Elbedour, May Weinberg, Gal Meiri, Analya Michaelovski, Idan Menashe

## Abstract

Maternal thyroid hormones play crucial roles in fetal neurodevelopment, with hypothyroidism and hyperthyroidism consistently being reported as potential risk factors for autism spectrum disorder (ASD) in offspring. Nevertheless, the mechanism underlying this association remains vague. In this retrospective cohort study of 51,296 singleton births in a single hospital, we examined the impact of different maternal thyroid conditions on the risk of ASD in the offspring. Chronic and gestational hyper-and hypothyroidism were determined based on ICD-9 criteria and maternal thyroid hormone levels, respectively. ASD diagnosis was determined based on DSM-5 criteria. Kaplan-Meier plots and Cox regression models were used to assess ASD risk in the offspring of women with and without thyroid conditions. A total of 4,306 (8.4%) of the mothers showed abnormal thyroid function. ASD cumulative incidence was similar in the offspring of women with normal and abnormal thyroid function (log-rank p=0.28). However, further classification of the study groups revealed an increased ASD risk in the offspring of women with both chronic and gestational thyroid dysfunction (aHR= 2.68, 95%CI=1.52-4.72). In addition, trimester-specific analysis revealed a dose effect in that the longer the period of thyroid imbalance, the higher the ASD risk, namely, for one, two or three trimesters of hypo- or hyperthyroidism, aHR= 1.69 (95%CI=1.19-2.83), aHR=2.39 (95%CI=1.24-5.78), aHR=3.25 (95%CI=1.07-7.21), respectively. Thus, the findings suggest that uncontrolled gestational thyroid dysfunction is associated with an elevated risk of ASD in offspring, underscoring the importance of routine thyroid function screening and treatment throughout pregnancy in mitigating ASD risk in offspring.

## Introduction

Autism spectrum disorder (ASD) refers to a range of neurodevelopmental conditions marked by the pervasive manifestation of deficits in social communication as well as by restricted and repetitive behaviors, interests, and activities [1]. ASD is characterized by a heterogeneity of phenotypic presentations, with its etiology lying in a range of genetic predispositions [2-8] in combination with pre- and post-natal environmental impacts [9-15]. Nonetheless, the exact pathways of ASD pathogenesis and its diagnostic markers remain elusive.

Ample research has indicated the contribution of the prenatal period to the development of ASD in offspring through the disturbance of critical neurodevelopmental pathways in the developing fetus. Particularly, a range of maternal risk factors have been associated with ASD risk in offspring [9, 15, 16-20]. A noteworthy prenatal risk that has been researched in the context of ASD is maternal thyroid dysfunction in pregnancy. Thyroid hormones, i.e., thyroid-stimulating hormone (TSH), triiodothyronine (T_3_), and thyroxine (T_4_), are essential for fetal neurogenesis and maturation, with the embryo/fetus being dependent on the supply of maternal thyroid hormones across the placenta, particularly in the first trimester [21].

Consequently, disruptions in maternal thyroid hormone levels during pregnancy have been associated with adverse developmental outcomes in offspring [21-23]. Maternal hypo- and hyperthyroxinemia have been associated with lower IQ scores and delays in expressive language and non-verbal cognition in children when compared to children born to mothers with adequate T_4_ levels [24-25]. Increased TSH (indicative of overt and subclinical hypothyroidism) has also been associated with poorer child cognitive function [26-27]. In addition, it has been suggested that autoimmune processes, which often play a role in both overt and subclinical hypothyroid etiology, independently impact fetal neurodevelopment beyond the influence of maternal thyroid dysfunction [28].

Previous research has also indicated that maternal thyroid dysfunction during pregnancy can alter fetal brain development, potentially increasing the likelihood of ASD. Some studies have found that untreated maternal hypothyroidism during pregnancy is associated with an increased risk of ASD in the offspring [29-30]. Other, smaller, studies investigating various maternal thyroid conditions have supplied supporting evidence for this association [31]. Although gestational screening and therapy of the relevant thyroid anomalies were recommended in those studies, medication use was not directly investigated. Another study of importance examined the association between maternal thyroid disorders and ASD likelihood in offspring registered with a large Israeli health fund [32]: it showed that children of mothers who had been diagnosed with hypo- or hyperthyroidism at some stage in their lives had higher odds of developing ASD compared to children of mothers with no such diagnosis [32]. Interestingly, the authors of that study suggested that the mechanism underlying the observed association was not related to the alterations in maternal gestational thyroid hormone concentrations, but rather, it could be attributed to a confounder that was independently associated with increased odds of both maternal thyroid dysfunction and ASD [32]. Thus, the lack of clarity and consistency in different research methods and conclusions attests to the many questions in this field that remain unanswered.

Thyroid disorders are widespread among women of reproductive age, and readily accessible diagnostic methods allow for a high prevalence of treatment in this demographic stratum [33]. Nonetheless, thyroid hormone levels tend to fluctuate intermittently, and stabilization of hormone levels may prove difficult, particularly during pregnancy [33]. Thus, to accurately assess the association between maternal thyroid disease state and offspring ASD, we chose to determine maternal thyroid dysfunction on the basis of clinically measurable blood test results, irrespective of a diagnosis of thyroid dysfunction according to the International Classification of Diseases, Nineth Revision (ICD-9). Consequently, our methodology enables us to distinguish between women diagnosed with a chronic thyroid condition (likely being controlled by thyroid medications) and women with an established thyroid hormone imbalance during pregnancy.

## Methods

### Study population

We conducted a retrospective cohort study of live singleton births at Soroka University Medical Center (SUMC) between January 2011 and December 2017 to mothers who were members of Clalit Health Services (CHS), the largest health maintenance organization in Israel. SUMC serves as the sole tertiary medical center in southern Israel and is a referral center for patients with mild to complex conditions from the entire Negev region. The population of the Negev region consists of approximately 1 million citizens, of which over 75% are members of the CHS. Furthermore, the Negev population has a unique ethnic composition in that more than 25% of its inhabitants are Bedouins. Importantly, >50% of the births at SUMC are births for this population sector.

### Data collection

We used mothers’ ID numbers to cross-link and merge data from three medical resources: (1) the CHS electronic database, which included sociodemographic information and data on chronic diagnoses for all women in the study; (2) the database of the Obstetrics and Gynecology Department at SUMC, which contains comprehensive prenatal and perinatal data on all women giving birth at SUMC since 1990; and the database of the Azrieli National Center for Autism and Neurodevelopment Research (ANCAN), which contains data on all children who have been diagnosed with ASD according to DSM-5 criteria [1] at SUMC since 2013 [34-35].

### Maternal thyroid dysfunction

We identified women with chronic, gestational, or both chronic and gestational thyroid conditions as follows. Diagnoses of chronic hypothyroidism or hyperthyroidism were made according to ICD-9 [36], as recorded in the CHS database. Data on gestational thyroid hormones levels that are routinely measured throughout pregnancy were obtained from the SUMC database. Women without any test of thyroid hormones during pregnancy were excluded from the study. We used the median of each woman’s thyroid hormone values in each pregnancy trimester to identify one of three types of thyroid dysfunction in each trimester. *Gestational hypothyroidism* was defined as a median T_4_ < 0.8 ng/dL and a median TSH > 4 IU/mL (overt hypothyroidism) or a median T_4_ = 0.8–1.5 ng/dL and a median TSH > 4 IU/mL (subclinical hypothyroidism). *Gestational hyperthyroidism* was defined as a median T_4_ > 1.5 ng/dL and a median TSH < 4 IU/mL. A woman was defined as having gestational hypothyroidism or hyperthyroidism if she met the above criteria in one or more of her pregnancy trimesters. Finally, women were classified as having *chronic thyroid dysfunction*, gestational thyroid dysfunction, or both. A flowchart depicting the study sample ascertainment is provided in Fig. 1.

**Fig. 1.**
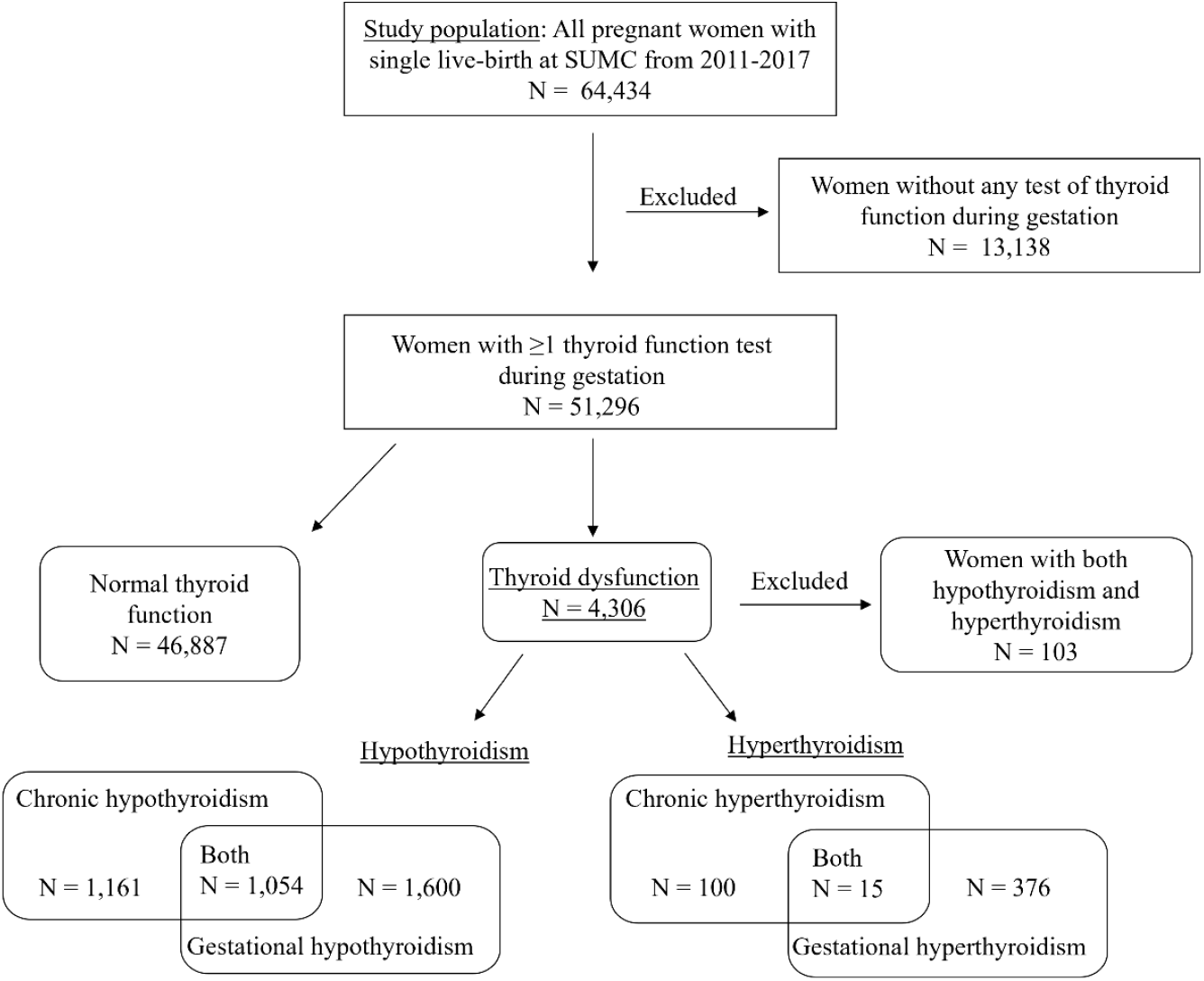
Flowchart depicting ascertainment of the study groups.

### Data analysis

Mean and standard deviation (SD) values were calculated for continuous variables, while percentages were calculated for nominal variables. Univariate analysis was performed by either two-tailed t-tests or Mann-Whitney U tests for continuous variables, and two-sided *χ2* tests for nominal variables. Cox regression models were performed to assess both the hazard associated with exposure to the various thyroid dysfunction states, as well as the exposure across different trimesters, while adjusting for confounding. All statistical analyses were performed with SPSS version 28.0. Statistically significant results were determined at p < 0.05.

## Results

Of the 51,296 women included in the study, there were 4,306 (8.4%) with abnormal thyroid function. Of these, 1,161 had chronic hypothyroidism, 1,600 had gestational hypothyroidism, and 1,054 had both chronic and gestational hypothyroidism; in addition, 100 women had chronic hyperthyroidism, 376 had gestational hyperthyroidism, and 15 women had both these conditions (Fig. 1). Notably, for 103 women there were indications of both hypothyroidism and hyperthyroidism (either chronic or gestational; supplementary Fig. S1), and these women were therefore excluded from further analysis (Fig. 1).

Socio-demographic and clinical characteristics of the study sample, divided into women with normal thyroid function and with any thyroid dysfunction, are detailed in Table 1. The study groups varied significantly in their ethnic composition (Jewish vs. Bedouin), with 43.3% and 49.6% of the women in normal thyroid function and with any thyroid dysfunction groups being Jewish (p < 0.001). Additional differences between women with normal thyroid function vs. those with any thyroid dysfunction were seen in maternal age (28.97 ± 5.75 vs. 29.93 ± 5.95 years, p < 0.001), rates of assisted reproductive therapy (4.4% vs. 6.5%, p < 0.001), and rates of in-vitro fertilization (0.3% vs. 0.5%; p = 0.023), as well as in various prenatal and perinatal conditions, such as gestational diabetes (6.8% vs. 9.5%, p < 0.001), hypertension (9.1% vs. 10.9%, p < 0.001), Cesarean section (25.8% vs. 29.2%, p < 0.001) and assisted delivery (36.2% vs. 39.4%, p < 0.001). Regarding offspring characteristics, there was a slight overrepresentation of male children in the total sample without significant differences between the above two groups (52.4% and 52.2%, respectively). The offspring of the two groups also did not differ in their birth weight or gestational age.

**Table 1.**
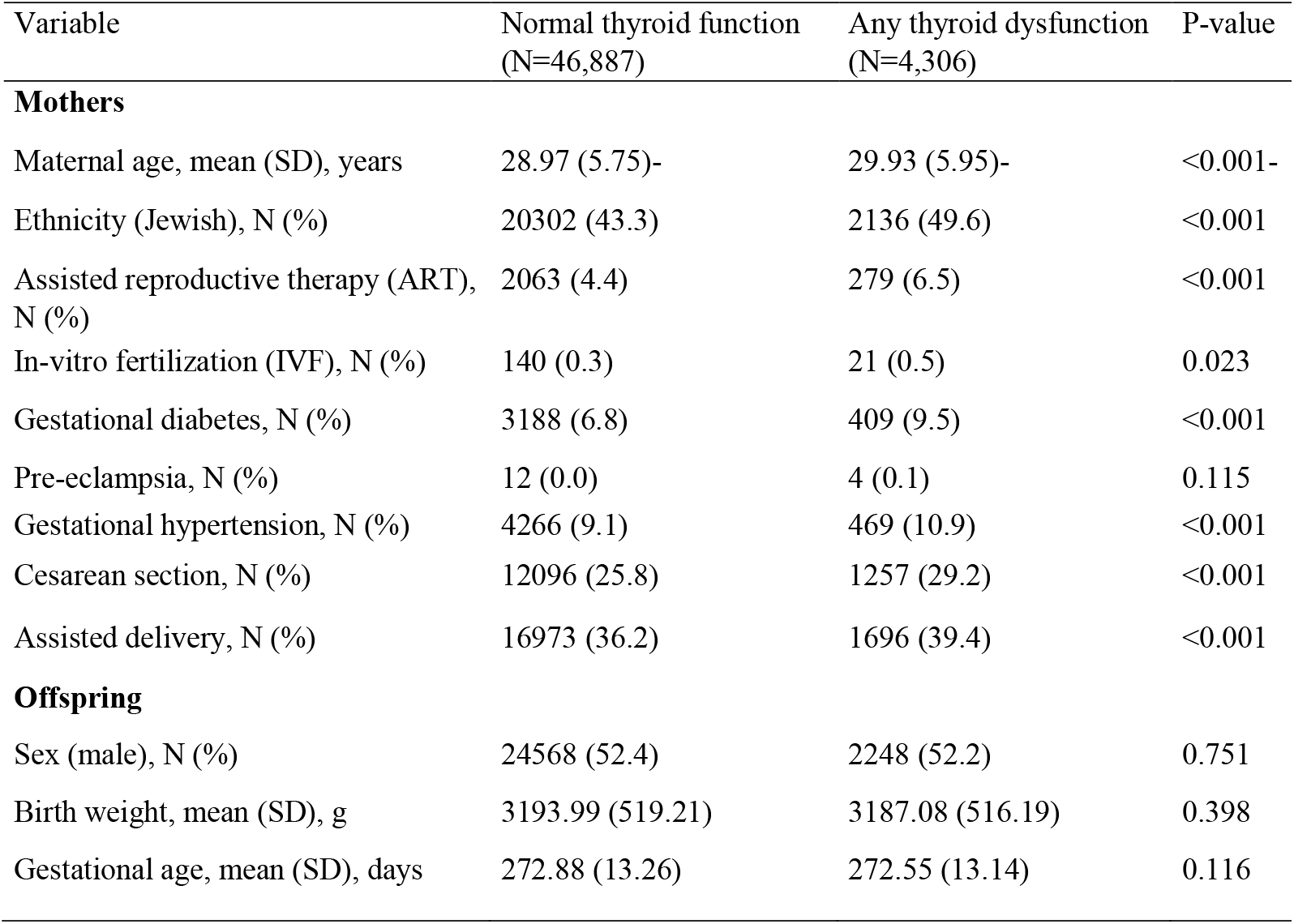
Socio-demographic and clinical characteristics of study sample.

| Variable | Normal thyroid function<br>(N=46,887) | Any thyroid dysfunction<br>(N=4,306) | P-value |
| --- | --- | --- | --- |
| **Mothers** |  |  |  |
| Maternal age, mean (SD), years | 28.97 (5.75)- | 29.93 (5.95)- | <0.001- |
| Ethnicity (Jewish), N (%) | 20302 (43.3) | 2136 (49.6) | <0.001 |
| Assisted reproductive therapy (ART),<br>N (%) | 2063 (4.4) | 279 (6.5) | <0.001 |
| In-vitro fertilization (IVF), N (%) | 140 (0.3) | 21 (0.5) | 0.023 |
| Gestational diabetes, N (%) | 3188 (6.8) | 409 (9.5) | <0.001 |
| Pre-eclampsia, N (%) | 12 (0.0) | 4 (0.1) | 0.115 |
| Gestational hypertension, N (%) | 4266 (9.1) | 469 (10.9) | <0.001 |
| Cesarean section, N (%) | 12096 (25.8) | 1257 (29.2) | <0.001 |
| Assisted delivery, N (%) | 16973 (36.2) | 1696 (39.4) | <0.001 |
| **Offspring** |  |  |  |
| Sex (male), N (%) | 24568 (52.4) | 2248 (52.2) | 0.751 |
| Birth weight, mean (SD), g | 3193.99 (519.21) | 3187.08 (516.19) | 0.398 |
| Gestational age, mean (SD), days | 272.88 (13.26) | 272.55 (13.14) | 0.116 |

Cumulative incidence of ASD in the offspring of the women of the two study groups (normal vs. any thyroid dysfunction) is depicted in Fig. 2. No significant differences in ASD cumulative incidence was observed between offspring of women with normal and abnormal thyroid function (Fig. 2A; log-rank p = 0.28). However, further classification of the study groups into gestational and chronic thyroid dysfunction revealed a significantly higher cumulative ASD incidence in children born to mothers diagnosed with both chronic and gestational thyroid dysfunction when compared to those with normal thyroid function (Fig. 2B; log-rank test, p = 0.0002). Conversely, cumulative incidence of ASD in offspring of women with chronic thyroid dysfunction alone or gestational thyroid dysfunction alone did not differ from the cumulative incidence of ASD in offspring of women with normal thyroid function (Fig. 2B).

**Fig. 2.**
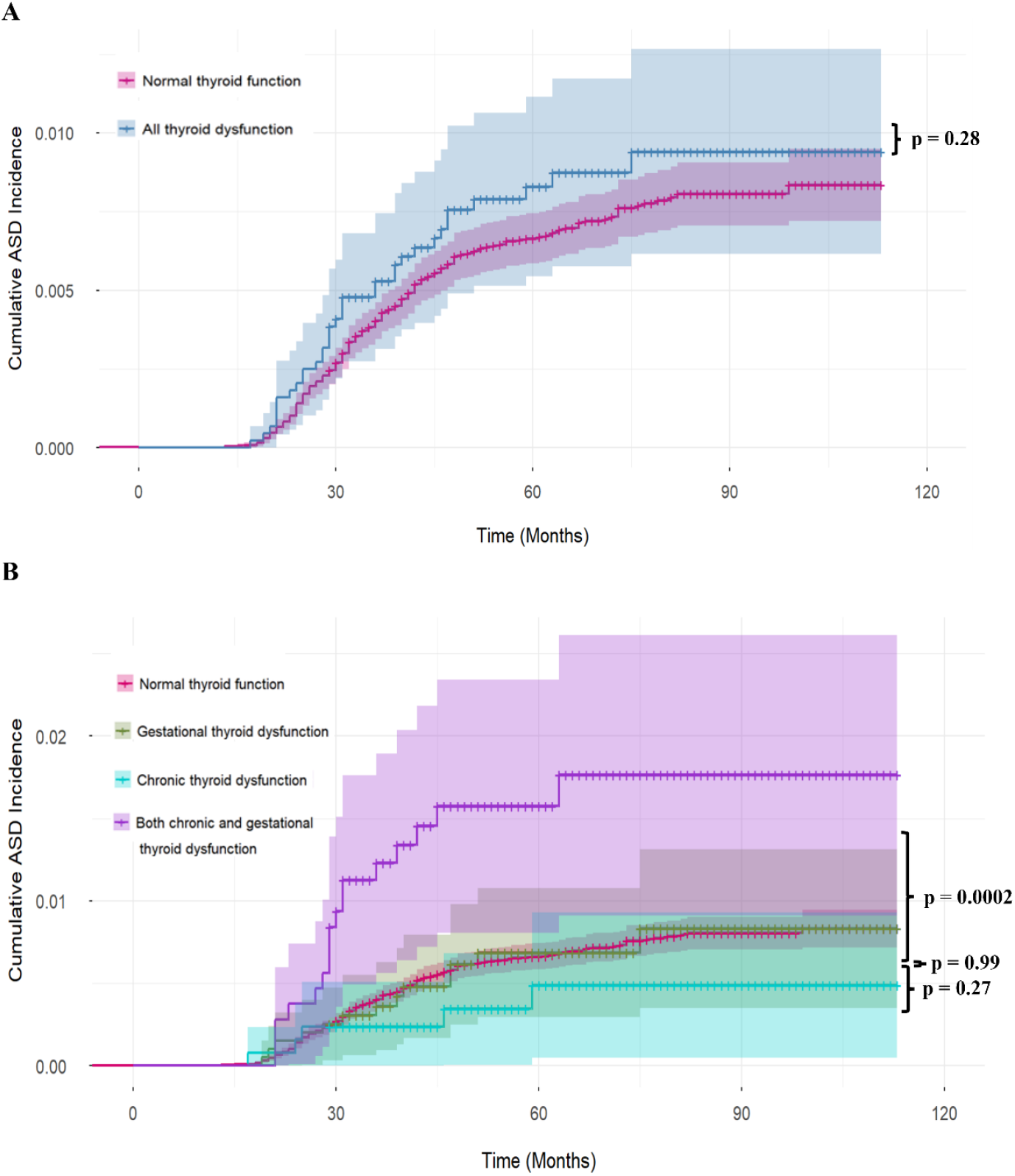
Kaplan-Meier plots for cumulative incidence of ASD onset in various thyroid function states. A) ASD incidence in women with normal thyroid function versus in those with overall thyroid dysfunction (both hypothyroidism and hyperthyroidism). B) ASD incidence in women with chronic, gestational, and both chronic and gestational thyroid dysfunction versus in those with normal thyroid function.

Cox regression models were used to assess the hazard ratio of various types of maternal thyroid dysfunction associated with an offspring diagnosis of ASD. The results of these analyses are presented in Table 2. There was no association between any type of thyroid dysfunction (hypothyroidism or hyperthyroidism during gestation or as a chronic condition) and ASD risk in either the crude or adjusted model. Further division of the thyroid dysfunction group into women with any type of hypothyroidism or any type of hyperthyroidism showed opposite trends (although not statistically significant), with a slightly higher risk of ASD in the offspring of women with hypothyroidism than of women with normal thyroid function (aHR=1.31; 95%CI = 0.83-2.07), while the offspring of women with hyperthyroidism had a lower risk of ASD (aHR=0.49; 95%CI = 0.07-3.53). Importantly, a division of the study groups according to a chronic vs. a gestational thyroid condition revealed an increased risk of a later diagnosis of ASD only in the offspring of women with both chronic and gestational thyroid dysfunction (aHR = 2.68, 95%CI = 1.52-4.72). This increased risk of ASD remained statistically significant separately in women with chronic and gestational hypothyroidism and women with chronic and gestational hyperthyroidism (aHR = 2.61, 95%CI = 1.44-4.74; aHR = 11.67, 95%CI = 1.29-105.83, respectively), suggesting that combined maternal chronic and gestational thyroid dysfunction, but neither of these two conditions alone, is a risk factor for ASD in the offspring. Further classification of gestational hypothyroidism into subclinical and overt hypothyroidism revealed a possible dose-response effect of thyroid hormone levels, in that offspring of women with the overt condition were at higher risk of ASD than those of women with the subclinical condition (aHR = 2.11, 95%CI = 0.86-5.18; aHR = 1.65, 95%CI = 1.10-2.48, respectively).

**Table 2.** Hazard ratio (HR) of thyroid conditions associated with ASD risk.

| Status | HR | 95% CI | Adjusted HR* | 95% CI |
| --- | --- | --- | --- | --- |
| Thyroid dysfunction<br>(N = 4306) | 1.31 | 0.84-2.04 | 1.18 | 0.75-1.84 |
| All Hypothyroidism<br>(N = 3815) | 1.47 | 0.93-2.31 | 1.31 | 0.83-2.07 |
| All Hyperthyroidism<br>(N = 491) | 0.52 | 0.07-3.69 | 0.49 | 0.07-3.53 |
| Chronic condition only<br>(N = 1261) | 0.56 | 0.18-1.75 | 0.43 | 0.14-1.35 |
| Gestational condition only<br>(N = 1976) | 0.82 | 0.37-1.86 | 0.87 | 0.39-1.96 |
| Chronic and gestational conditions<br>(N = 1069) | 3.03 | 1.73-5.31 | 2.68 | 1.52-4.72 |
| Chronic hypothyroidism only<br>(N = 1161) | 0.64 | 0.20-1.99 | 0.47 | 0.15-1.48 |
| Chronic hyperthyroidism only<br>(N = 100) | -- | -- | -- | -- |
| Gestational hypothyroidism only<br>(N = 1600) | 1.11 | 0.49-2.50 | 1.19 | 0.52-2.70 |
| Gestational hyperthyroidism only<br>(N = 376) | -- | -- | -- | -- |
| Chronic and gestational<br>hypothyroidism<br>(N = 1054) | 2.87 | 1.59-5.17 | 2.61 | 1.44-4.74 |
| Chronic and gestational<br>hyperthyroidism<br>(N = 15) | 19.78 | 2.49-156.91 | 11.67 | 1.29-105.83 |
| Subclinical hypothyroidism<br>(N = 2414) | 1.72 | 1.15-2.57 | 1.65 | 1.10-2.48 |
| Gestational overt hypothyroidism<br>(N = 388) | 2.01 | 0.83-4.88 | 2.11 | 0.86-5.18 |

\*Adjusted for sex, ethnicity, gestational age, maternal age, birth weight, assisted reproductive therapy, in-vitro fertilization, gestational diabetes, gestational hypertension, pre-eclampsia, Cesarean section, and assisted delivery.

Finally, we examined the association between maternal gestational hypothyroidism across different pregnancy trimesters and offspring ASD (Table 3). Once again, an interesting dose effect was observed in that a longer period of hypothyroidism during pregnancy was associated with a higher risk of ASD in the offspring. Specifically, the adjusted HR for ASD in the offspring for women with hypothyroidism in only one pregnancy trimester was 1.69 (95%CI = 1.19-2.83), while hypothyroidism in two trimesters increased the offspring ASD risk to aHR=2.39 (95%CI = 1.24-5.78), and hypothyroidism throughout the pregnancy further increased the risk to aHR=3.25 (95%CI = 1.07-7.21). Unfortunately, a similar analysis for gestational hyperthyroidism was not possible due to the small number of women with this condition.

**Table 3.** Hazard ratio (HR) of gestational hypothyroidism associated with in ASD across trimesters.

| Trimester | HR | 95% CI | Adjusted HR* | 95% CI |
| --- | --- | --- | --- | --- |
| 1st trimester only<br>(N = 571) | 1.71 | 1.14-3.09 | 1.82 | 1.22-3.21 |
| 2nd trimester only<br>(N = 432) | 1.43 | 0.95-3.43 | 1.39 | 0.91-3.22 |
| 3rd trimester only<br>(N = 246) | 1.88 | 1.30-4.17 | 1.84 | 1.21-4.09 |
| 1st and 2nd trimesters<br>(N = 189) | 2.61 | 1.73-5.22 | 2.59 | 1.79-5.34 |
| 2nd and 3rd trimesters<br>(N = 54) | 2.32 | 0.98-5.15 | 2.22 | 0.87-5.87 |
| 1st and 3rd trimesters<br>(N = 20) | 2.78 | 0.75-10.57 | 2.81 | 0.76-11.35 |
| All trimesters (1st, 2nd, and 3rd)<br>(N = 88) | 3.23 | 1.01-7.76 | 3.25 | 1.07-7.21 |
| All single trimesters combined<br>(N = 1249) | 1.73 | 1.17-2.87 | 1.69 | 1.19-2.83 |
| All double trimesters combined<br>(N = 263) | 2.66 | 1.39-5.57 | 2.39 | 1.24-5.78 |
| All trimesters<br>(N = 88) | 3.23 | 1.01-7.76 | 3.25 | 1.07-7.21 |

\*Adjusted for sex, ethnicity, gestational age, maternal age, birth weight, assisted reproductive therapy, in-vitro fertilization, gestational diabetes, gestational hypertension, pre-eclampsia, Cesarean section, and assisted delivery.

## Discussion

The results of this study suggest that the offspring of women with a thyroid hormone imbalance during gestation, but not with other, supposedly controlled, forms of chronic thyroid dysfunction, have an increased risk of ASD as compared to the offspring of women with normal thyroid function. Our results are in keeping with a growing body of evidence indicating that irregular maternal thyroid hormone levels during gestation are associated with various fetal neurodevelopmental outcomes, including ASD [30-31, 37-38]. Additional studies that support our findings include a large epidemiological study thar showed abnormal first-trimester thyroid hormone levels to be associated with child ASD risk in cases where the mothers had not previously been diagnosed with thyroid dysfunction [39]. Another study found that the likelihood of ASD was not increased in the children of women diagnosed with chronic hypothyroidism, but with normal TSH and T_4_ levels during pregnancy due to medication [40].

Our results further demonstrate that ASD risk increases in direct relation to the severity and duration of thyroid hormone imbalance during pregnancy. While several studies with smaller sample sizes have failed to establish clear patterns linking thyroid dysfunction severity to ASD risk [39, 41], other studies support our findings that both subclinical and overt hypothyroidism are associated with ASD, with overt hypothyroidism being associated with a higher risk of ASD [42]. Additionally, some studies have reported an inverse association between ASD-related traits and TSH levels in the second trimester [43-44], which was the trimester most weakly associated with ASD risk in our trimester-specific analysis.

Notably, our results are somewhat in disagreement with the conclusions of another study conducted in Israel [32]. The authors of that study suggested that the association between maternal thyroid dysfunction and ASD in the offspring is not directly driven by gestational thyroid imbalance, but rather by another factor underlying both gestational thyroid dysfunction and ASD [32]. The discrepancy between the two studies may be attributed to the different ways gestational thyroid dysfunction was determined: while we exclusively used blood levels of thyroid hormones (TSH and T_4_) during gestation to determine thyroid dysfunction status, Rotem et al. primarily used the ICD-9 classification and medication dispensing records [32]. Thus, they assumed that the levels of thyroid hormones of women with thyroid disease were balanced by medication—an assumption that was shown to be invalid for a considerable portion of women in the present study.

A clear strength of the present study lies in the availability both of data for laboratory-determined thyroid hormone levels throughout pregnancy and of data for chronic thyroid conditions for the women in our study sample. Consequently, these data enabled us to distinguish, with a high degree of certainty, between women with a thyroid hormone imbalance during pregnancy and women diagnosed with a chronic thyroid condition that would probably be treated with thyroid medications, thereby conferring normal thyroid hormone levels during pregnancy. In addition, our study is among the first to consider the association between maternal hypothyroidism and risk of ASD across all the trimesters of pregnancy. A large number of previous studies investigating the association between maternal thyroid hormone levels and offspring ASD risk have determined thyroid hormone levels only at a single time point during gestation, thereby limiting a comprehensive understanding of the exposure-outcome relationship.

The limitations of the current study include the lack of data on thyroid autoimmunity as a potential mediator of ASD risk, although Rotem et al. [32] reported that there was no association or effect modification in its inclusion. Another limitation of the study is the decreased power of the trimester-specific analysis, due to the small number of women with gestational hypothyroidism in each trimester. This may be because once thyroid dysfunction is identified and promptly treated, most women exhibit a positive response to the treatment within 6 weeks of treatment [45]. Finally, the low prevalence of gestational hyperthyroidism did not allow us to separately study its association with ASD risk in offspring.

## Conclusions

Our findings suggest that an imbalance of maternal thyroid hormone concentrations during gestation is associated with an increased risk of ASD in the offspring in a dose-response manner. Thus, careful monitoring of thyroid hormone levels throughout pregnancy and treatment of imbalanced thyroid function with thyroid hormone medications may mitigate the risk of ASD in offspring.

## Data Availability

All data produced in the present study are available upon reasonable request to the authors

## Acknowledgment

We thank Mrs. Inez Mureinik for critically reviewing and editing the manuscript

## Declarations

### Ethics approval

The research was prospectively reviewed and approved by the ethics committee of SUMC (#237-21).

**Supplementary Fig. S1.**
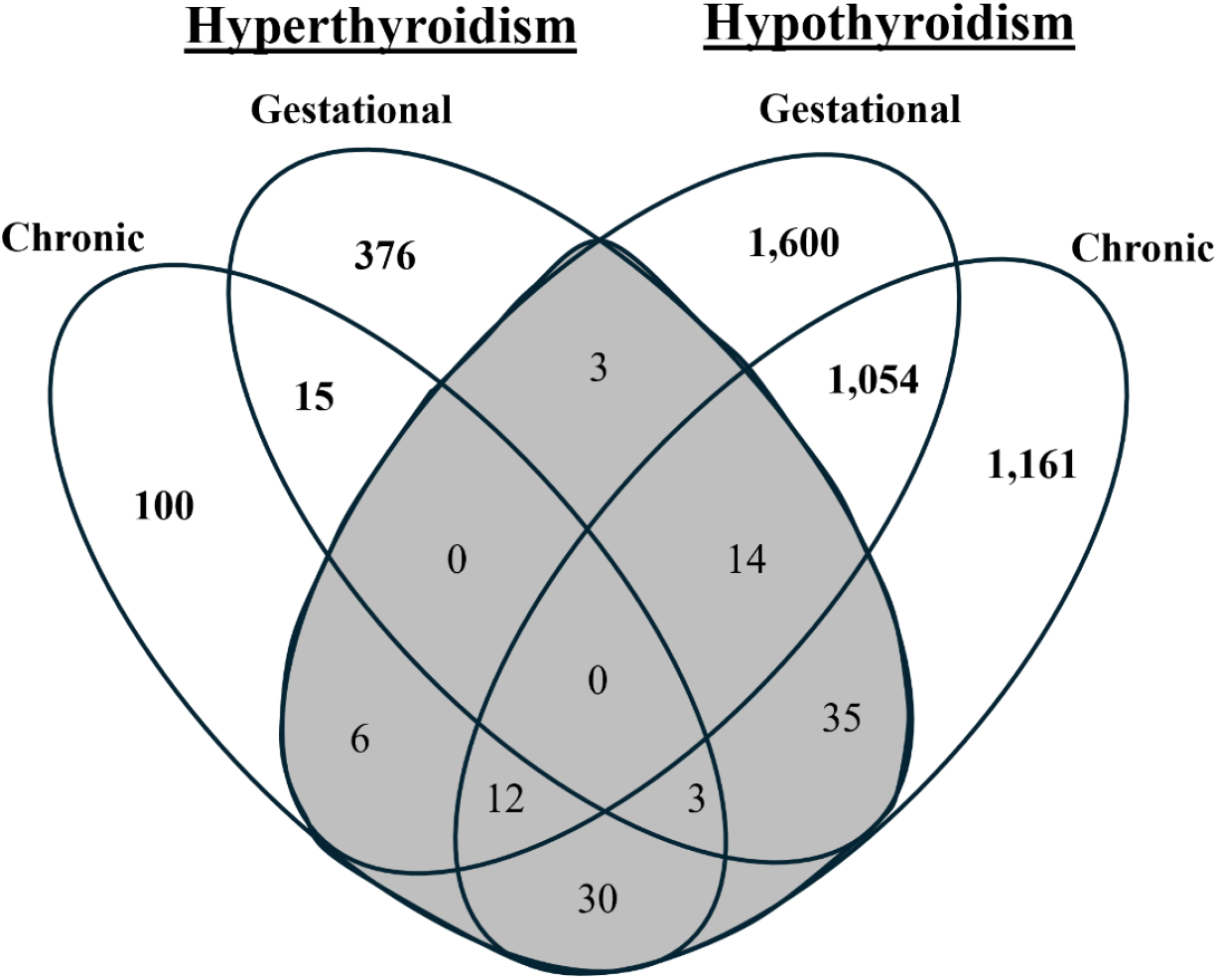
A Venn diagram depicting the different diagnoses of thyroid dysfunction in women in the study. The gray area depicts 103 women with indications of both hypothyroidism and hyperthyroidism during the study period.

